# Loss of chromosome Y shapes the immune cell fate with aging in men

**DOI:** 10.1101/2025.06.01.25328624

**Authors:** Ahmed Dawoud, Luke Green, Owen Rackham

## Abstract

The loss of chromosome Y (LOY) in leukocytes is the most prevalent form of clonal mosaicism observed in older men. It is associated with all major causes of mortality, including cardiovascular diseases and cancer. However, LOY effect on individual immune-cell lineages remains unclear.

Here, we used single-cell RNA sequencing data from Onek1K cohort and showed that LOY has a wide effect on cell fate across immune cell populations with the largest representation of LOY in classical monocytes. These monocytes exhibit a profibrotic signature marked by downregulation of *IL1B* and MYC-regulated genes, consistent with previous observations of LOY-associated macrophages in cardiac and pulmonary injury. Additionally, we detected an aberrant expression of *XIST*, the essential X-chromosome inactivation (XCI) lncRNA expressed in females, and not normally expressed in males. Notably, we observed upregulation of genes known to escape X-inactivation, including male-biased cancer-related genes *KDM6A*, *DDX3X*, *KDM5C, and ZRSR2*.

Our results demonstrate the effect of LOY on the fate of immune cells, mediated by cell-type specific transcriptional changes and aberrant XCI characteristics.

## Main

Human haematopoiesis is the efficient, yet age-dependent, process of generating blood and immune cells. This process is challenged by acquisition of somatic point mutations^1,2^ and/or large chromosomal alterations^3,4^ over time. These mutations can provide a selective growth advantage and drive clonal expansion in blood ^5^. Among all detected somatic mutations, the loss of chromosome Y (LOY) in the peripheral blood of men is the most prevalent event^6,7^ that affects more than half of men by age of 80 years^8,9^. The Y chromosome is the smallest human chromosome and plays a key role in male sex determination, hosting genes essential for spermatogenesis^10^. However, large population studies have indicated a wider role for Y chromosome in the progression of haematological malignancies^11^, solid cancers ^6^, non-malignant diseases such as Alzheimer’s disease^12^ , and cardiovascular diseases^13,14^.

One common feature of these disease associations is the effect of LOY on the immune system, either at the population scale or in specific disease contexts. For example, LOY has been associated with an increase in leukocytes in Biobank Japan cohort^15^ and more specifically neutrophils and monocytes in UK Biobank cohort^16^. An association between LOY and an increase in Natural killer (NK) cells was observed in Alzheimer’s disease^17^, while LOY led to an increase in CD4+ T cells in prostate cancer^17^ and pulmonary fibrosis^18^. Beyond these associations, LOY has also been shown to alter immune cell fates. LOY promoted a bias towards regulatory-T-cell-fate within the CD4+ population in peripheral blood and tissues^19^. Similarly, LOY was shown to promote a bias toward fibrotic rather than inflammatory phenotypes in monocyte-derived cardiac^13^ and alveolar^18^ macrophages. Collectively, these associations between LOY and the immune system highlight the potential value of LOY-associated markers as a novel therapeutic target, and risk predictors for age-related diseases^20^.

Genome-wide association studies (GWAS) had revealed the genetic risk factors of LOY in healthy individuals^7,9,15,21^. The most prominent marker was rs2887399 located in the promoter region of *TCL1A*, a marker for B lymphocytes, where T allele was associated with lower risk of LOY^7^. Single cell RNA sequencing (scRNA-seq) technology has allowed the exploration of cell-type specific expression of the markers identified by GWAS^17^. *TCL1A* was up regulated 1.75 folds in B lymphocytes with LOY in comparison to non-LOY cells^9^. Furthermore, differential gene expression analysis of scRNA-seq data identified autosomal and X-linked genes associated with LOY^17,19,22^. These genes were enriched in pathways related to RNA processing, immune response, and cellular proliferation^17^. Further exploration of larger single cell datasets would help to characterize the effect of these associations on cell fate.

Onek1k^23^, part of Phase 1 of the Tenk10k, is an initiative that performed genotyping and transcriptional profiling of the peripheral blood mononuclear cells (PBMCs) of 982 to identify genetic risk factors associated with gene expression in a cell type level. In this study, we aimed to characterize the prevalence of LOY across major immune cell types and along their lineage developmental trajectories in individuals from Onek1k. Furthermore, we sought to identify autosomal and X-linked genes that are; associated with LOY, affect immune cell fate, and represent a potential therapeutic and diagnostic target for age-related diseases.

## Results

### Prevalence of LOY across different immune cell types

Recent advances in the multiplexing of scRNA-seq to handle cells derived from 100s of donors in a single experiment^24^ has powered a new range of applications including longitudinal studies, and genotype-expression associations^25^. We analysed the data from two independent methods; scRNA-seq of PBMCs, and SNP arrays of whole blood DNA, to call LOY in 416 males aged between 19 an 93 years (median=68) within the Onek1k cohort.

For calling LOY in scRNA-seq, we followed the previously published method^19^; LOY identity was allocated to a cell if no reads assigned to male specific region (MSY) region on Y chromosome using two methods for reads counting; cell-ranger^26^ and velocyto^27^. From a total of 517,412 cells, we identified 45,304 cells (8.76%) in 416 male donors with LOY (Supplementary Table 1). The prevalence of LOY cells at the individual level ranged from between 1% (n=10/1,123 cells) to 54% (n=1,416/2,621 cells) per donor. Expanded clonality, which we defined by the observation of LOY in 10% or more of cells, was assigned to 25.7% (n=107 donors, median age = 71). Adjusting for the effect of sequencing pool and other major sources of variation (see methods), the prevalence of LOY cells was significantly associated with age (β=0.013[0.009-0.016], *P*=8.35×10^-14^) (Fig. 1a).

**Fig. 1:**
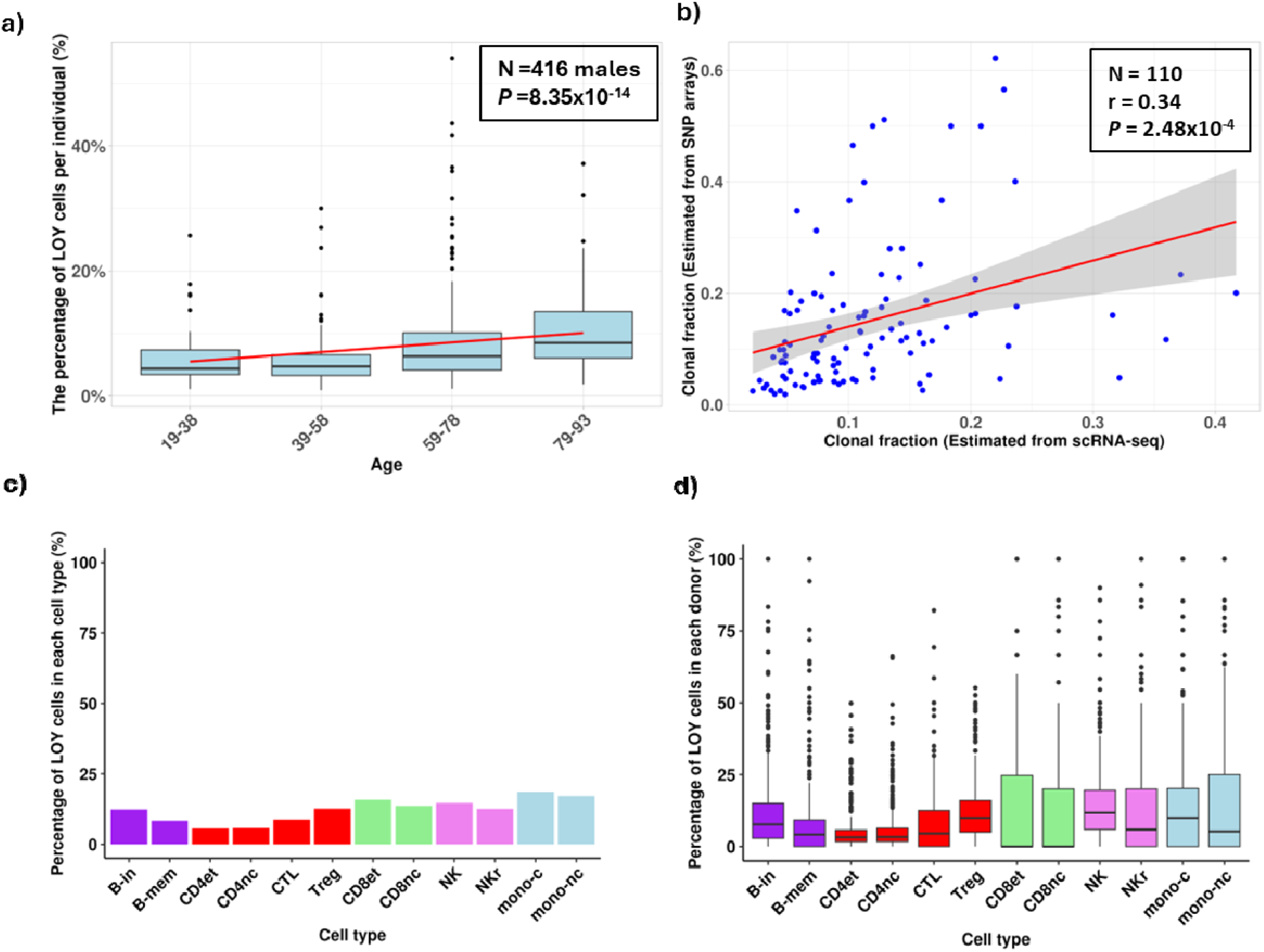
the prevalence of LOY in scRNA-seq and SNP arrays of males in Tenk10k data (a) Box plot illustrating the relationship between age, divided into four bins (19–38, 39–58, 59–78, and ≥79), and the percentage of LOY cells per individual. LOY was defined by the absence of expression counts of male-specific Y chromosome genes, derived from the combined Cell Ranger and Velocyto methods. The red line represents the linear regression fit. (b) Scatter plot showing the correlation between clonal fraction estimates from SNP arrays and single-cell data. Each dot represents an individual, and the red line represents the linear regression fit with a 95% confidence interval. (c) Bar plot showing the percentage of LOY cells identified in each cell type. (d) Box plot displaying the percentage of LOY cells per donor across different cell types. Dots indicate outliers.

For calling LOY in SNP-array data, we used phased-genotypes segmentation method^28^, and identified LOY in the leukocytes of 26.4% (n= 110) males with clonal fraction ranges between 0.02 and 0.62, and expanded clonality attribute in 58 males (13.9%). Notably, autosomal mosaic chromosomal alterations (mCA) were identified by SNP arrays data in 26 donors (28 events in total, Supplementary Table 1). Of them, 7 donors have both LOY and autosomal mCA. One important feature of PBMCs processed for scRNA-seq, in contrast to whole blood samples used for SNP arrays^29^, is that they only contain agranulocytes (monocytes and lymphocytes) and lack granulocytes (neutrophils, basophils, and eosinophils). However, genotyping data generated by SNP arrays and DNA extracted from whole blood samples is commonly used to assess acquired chromosomal mosaicism with sensitivity to 1% ^9,28^. We hypothesized that a significant proportion of LOY events with over representation in agranulocytes could be missed when we use DNA genotyping methods. To investigate this hypothesis, we assessed the relationship between the frequency of LOY per donor between scRNA-seq calls and SNP arrays calls. LOY identified by scRNA-seq has weak linear correlation with the clonal fraction detected by SNP arrays (r = 0.34; 95%CI, 0.17-0.50; *P*= 2.48×10^-4^, Fig. 1b). Furthermore, all males with expanded clonality, defined by clonal percentage 10% or more in SNP arrays data have been detected using scRNA-seq with 4.7% or more of LOY in PBMC cells, however only 50.5% (n=54/107) of expanded clonality defined by scRNA-seq calls were defined as LOY by SNP arrays. This observation suggests that SNP arrays failed to detect LOY events in leukocytes of donors with a high representation of LOY in the agranulocyte compartment. This highlights the potential of scRNA-seq to provide new insights into LOY, surpassing the limitations of genotyping-based methods

To assess the distribution of LOY across major cell type; monocytes, B-cells, Natural Killer cells, CD4+ T-cells, and CD8+ T-cells, we classified cells using a scheme derived from Yazar et al ^23^. In brief, we group cells into one of the five major cell lines, after excluding minor cell types. Each one of the major cell types were analysed independently, cells were clustered and annotated following the scheme in Table 1.

**Table 1:**
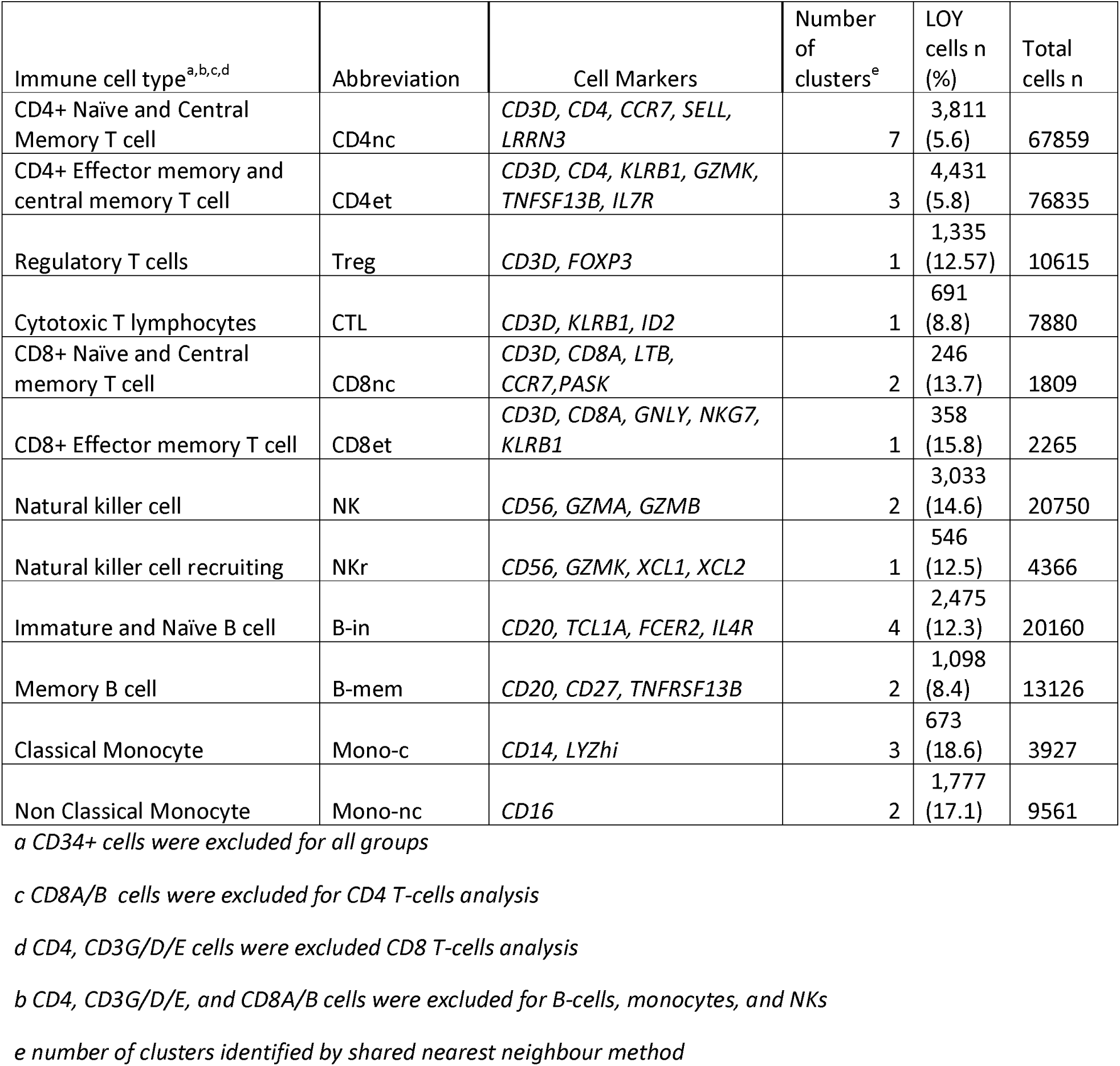
major cell types analysed for the prevalence of LOY across 416 donors.

| Immune cell type<sup>a,b,c,d</sup> | Abbreviation | Cell Markers | Number of clusters<sup>e</sup> | LOY cells n (%) | Total cells n |
| --- | --- | --- | --- | --- | --- |
| CD4+ Naïve and Central Memory T cell | CD4nc | *CD3D, CD4, CCR7, SELL, LRRN3* | 7 | 3,811 (5.6) | 67859 |
| CD4+ Effector memory and central memory T cell | CD4et | *CD3D, CD4, KLRB1, GZMK, TNFSF13B, IL7R* | 3 | 4,431 (5.8) | 76835 |
| Regulatory T cells | Treg | *CD3D, FOXP3* | 1 | 1,335 (12.57) | 10615 |
| Cytotoxic T lymphocytes | CTL | *CD3D, KLRB1, ID2* | 1 | 691 (8.8) | 7880 |
| CD8+ Naïve and Central memory T cell | CD8nc | *CD3D, CD8A, LTB, CCR7,PASK* | 2 | 246 (13.7) | 1809 |
| CD8+ Effector memory T cell | CD8et | *CD3D, CD8A, GNLY, NKG7, KLRB1* | 1 | 358 (15.8) | 2265 |
| Natural killer cell | NK | *CD56, GZMA, GZMB* | 2 | 3,033 (14.6) | 20750 |
| Natural killer cell recruiting | NKr | *CD56, GZMK, XCL1, XCL2* | 1 | 546 (12.5) | 4366 |
| Immature and Naïve B cell | B-in | *CD20, TCL1A, FCER2, IL4R* | 4 | 2,475 (12.3) | 20160 |
| Memory B cell | B-mem | *CD20, CD27, TNFRSF13B* | 2 | 1,098 (8.4) | 13126 |
| Classical Monocyte | Mono-c | *CD14, LYZhi* | 3 | 673 (18.6) | 3927 |
| Non Classical Monocyte | Mono-nc | *CD16* | 2 | 1,777 (17.1) | 9561 |

*a CD34+ cells were excluded for all groups*
*c CD8A/B cells were excluded for CD4 T-cells analysis*
*d CD4, CD3G/D/E cells were excluded CD8 T-cells analysis*
*b CD4, CD3G/D/E, and CD8A/B cells were excluded for B-cells, monocytes, and NKs*
*e number of clusters identified by shared nearest neighbour method*

Previous studies had shown the highest frequency of LOY in monocytes and NK ^17,19,22^, we refined this observation into defining classical monocytes (mono-c) to have the highest frequency (18.6% ) in all the study (Fig. 1c), and defining NK with the highest median of LOY per donor (11.76%, IQR=13.6) followed by Treg (10%, IQR=11) and mono-c (10%, IQR=20.2, Fig. 1d). The relationship between single cell calls and SNP arrays calls varies across cell types (Supplementary Table 2, and Supplementary Fig. 1). Clonal fraction estimated from SNP arrays has strong linear relationship with those estimated from scRNA-seq calls in monocytes subclasses; classical monocytes (mono-c ,r = 0.75, *P*_fdr_= 2.20×10^-11^), and non-classical monocytes (mono-nc, r = 0.66, *P*_fdr_ = 1.04 x 10^-5^). Moderate relationships were detected between the clonal fraction estimated from SNP arrays and those estimated from NK cells (NK, r = 0.51, *P*_fdr_ = 1.88 x 10^-7^), and NK recruiting cells (NKr, r = 0.57, *P*_fdr_ = 1.26 x 10^-4^), and weak correlation was identified with one subset of B cells; immature and naïve cells (B-in, r = 0.31, *P*_fdr_ = 4.59×10^-3^), but not with memory B cells (B-mem). However, we found no correlation between the clonal fraction estimated by SNP arrays and any of CD4 or CD8 T cells subtypes (i.e. naive and central memory (CD4nc), effector (CD4et), cytotoxic (CTL), and regulatory cells (Treg), naive and central memory (CD8nc), and effector memory (CD8et).

These observations highlight the bias of LOY calls derived from SNP arrays data toward presenting the myeloid compartment of immune cells, whereas LOY calls derived from scRNA-seq data were capable to assess both the myeloid and lymphoid compartments in a way that DNA genotypic methods cannot. In the following sections, we dissected the effect of LOY defined by scRNA-seq on the phenotype and transcriptional profiles of major immune cell types

### The effect of LOY on immune cells fate

Two independent studies have identified a lineage bias associated with LOY in CD4+ T cells, marked by an overrepresentation of LOY cells in T regulatory (Treg) cell populations within both peripheral blood and cancer tissues^19,30^. To further investigate potential lineage biases across major cell types, we assessed the relationship between LOY frequency and cell classification using two approaches: (i) by marker-defined subtypes (Fig. 2a, c, e, g, and i). and (ii) by the cell position on a cell-type developmental trajectory. These trajectories capture critical aspects of cell type flux (i.e. Immature and Naïve B cell transition to Memory B cell) and were stratified into six equal quantiles based on pseudotime along the trajectory (Fig. 2b, d, f, h, and j).

**Fig. 2:**
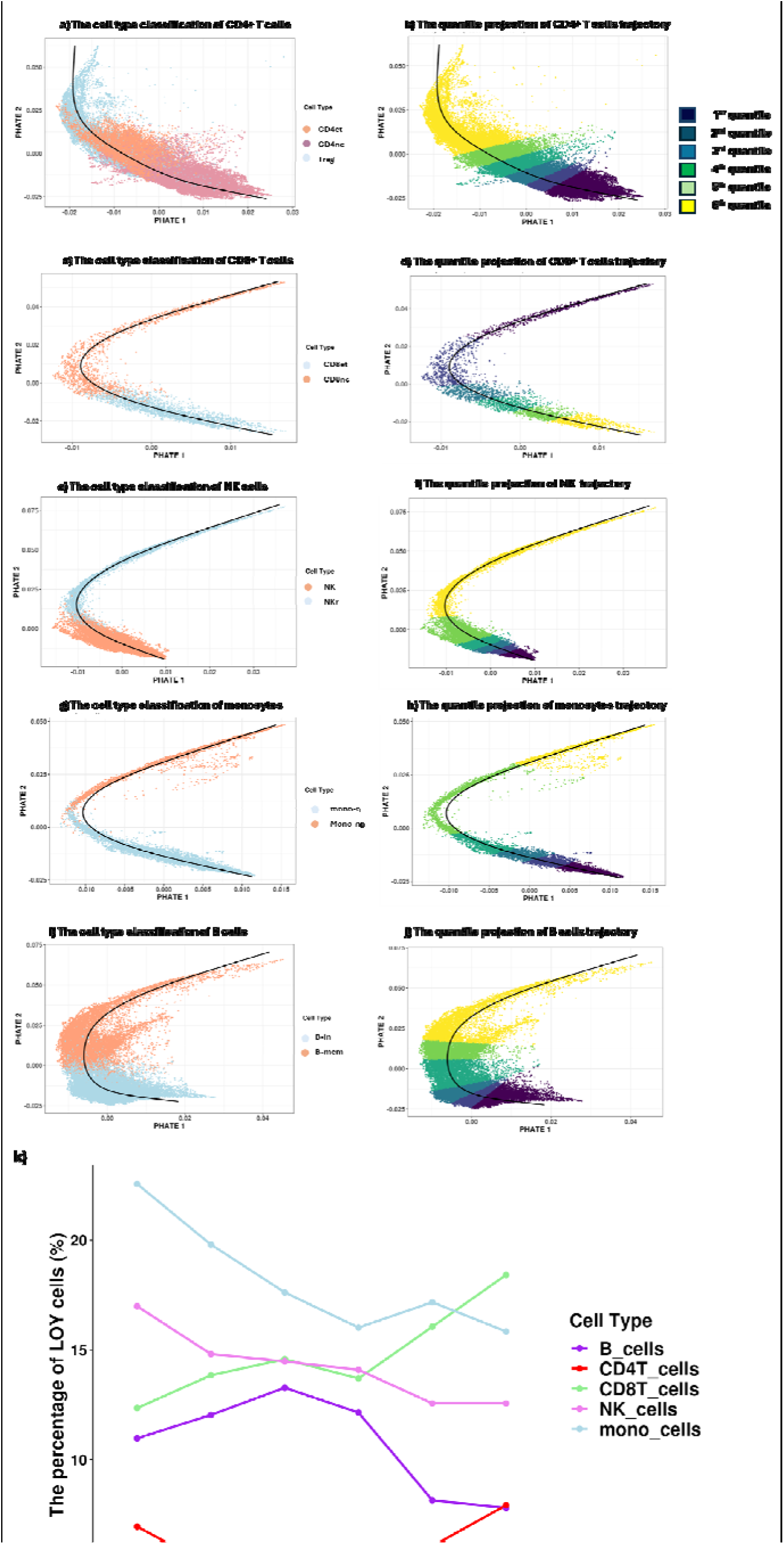
Cells were categorized into five major cell types and further classified along their developmental trajectory using pseudotime analysis, and six equal quantiles. (a,c,e,g,and i) Quantile projections visualized using PHATE dimensionality reduction. (b,d,f,h, and j) cell type projections visualized using PHATE dimensionality reduction. (a,b) Transition from immature and naïve B cells to B memory cells. (c,d) Transition from natural killer (NK) cells to NK recruiting cells. (e,f) Transition from CD8+ naïve and central memory T cells to CD8+ effector memory T cells. (g,h) Transition from classical monocytes to non-classical monocytes. (I,j) Transition from CD4+ naïve and central memory T cells to CD4+ effector memory and central memory T cells. (k) Line plot showing the relationship between the percentage of LOY cells and pseudotime quantiles across different cell types.

Initially, we confirmed the previous observation that LOY cells are more enriched in Treg cells (n=1335/10615) 12.57% in comparison to other CD4+ T cells; CD4nc (n=3811/67859) 5.6%, CD4et (n=4431/76835) 5.8%, and CTL (n=691/7880) 8.8%, and using a multivariate model adjusted for age, and first 10 genotypic principle components (PCs), (OR = 2.41, *P* = 2.28X10^-157^), (Supplementary Table 3). Next, we constructed the developmental trajectory of CD4+ T cells transitioning toward Treg (Fig2. a). The developmental trajectory of Treg cells represent their progression from the uncommitted naiive cells^31^. Notably, effector T cells can acquire regulatory functions under specific conditions, transitioning into Treg cells, marked by the expression of *FOXP3*^32^. We used PHATE dimensionality reduction to visualize the CD4+ T to T-reg manifold, and to construct its developmental trajectory. (see methods for details). This allowed to detect the expected positive correlation between LOY (OR=1.04, *P_adj_* = 6.92×10^-8^) and the positioning of cells along the developmental trajectory (Fig2. b,k). Interestingly, CD8+ T cells showed a gradual increase in LOY cells across the trajectory of differentiation (Fig2. d) from CD8nc (246/1809, 13.7%) to CD8et (358/2265, 15.8%) and from 12.4% in first quantile to a maximum of 18.4% in sixth quantile (OR=1.08, *P_adj_*=4.02×10^-3^).

In contrast, NK, monocytes, and B cells, showed a gradual decrease in the percentage of LOY cells along their development trajectory. NK cells undergo transformation along their development trajectory to activated state (Fig.2 e). In the activated state, the cells express chemokines such as *XCL1/2* to recruit other immune cells^33^. We referred to this state as NK recruiting (NKr) cells. LOY was less prevalent in NKr cells (546/ 4366, 12.5%) in comparison to other NK cells (3033/ 20750, 14.6%, OR=0.85, *P_adj_* = 2.03×10^-3^) with consistent decrease across the development trajectory from 17% in first quartile to 12.6% in the sixth quantile (Fig2. f).

Monocytes are the sole agranulocyte among myeloid cell types. An increased monocyte count is a characteristic feature of LOY^16^. CD14+ CD16− monocytes, known as classical monocytes (mono-c), represent 85% of the circulating monocytes, that emerge from bone marrow^34^. They play an essential role in inflammation by migrating to tissues and differentiating into macrophages or dendritic cells. In the circulating blood, classical monocytes transform into CD14+ CD16+ intermediate and CD14-CD16+ non-classical monocytes (mono-nc). To characterize the relationship between LOY and monocytes transition state, we constructed monocytes developmental trajectory to reflect the transition from mono-c to mono-nc (Fig.2 g). Initially, we detected a higher representation of LOY in mono-c (n=673/3927, 18.6%), against mono-nc (n=1777/9561, 17.1%, OR= 0.87, *P_adj_* = 0.02). This difference is consistent across the trajectory of monocytes (Fig2. h) and ranges between 22.6% in first quantile to 15.8% in sixth quantile (OR=0.91, *P_adj_* =1.71×10^-11^). This gradual decrease is remarkable in comparison to the short life span of monocytes that ranges between 1 day to 7 days^34^.

B cells are a component of the adaptive immune system, where immature and naiive B cells (B-in) differentiate into plasma cells, or memory B cells upon activation by antigens^35^. B cells produce antibodies of different immunoglobulin isotypes, whereas B-mem represent the secondary immune responses upon re-encountering the same antigen^36^. CD20 is a marker for B cells, that is downregulated upon differentiation to plasma cells. Our analysis was restricted to CD20+ clusters, and further classified these into B-in or B-mem as mentioned above, and using markers including *TCL1A* for B-in and *CD27* for B-mem (Fig.2 i) . Here, we observed a decrease in LOY cells from (n=2475/20160,12.3%) in B-in to (n=1098/13126, 8.4%, OR= 0.63, *P_adj_* = 4.88×10^-20^) in B-mem. Furthermore, we identified a gradual decrease in LOY cells on the transition trajectory (Fig2. j) from B-in to B-mem (OR=0.92, *P_adj_* = 7.30×10^-12^).

*TCL1A* is a marker for B-in that down regulated gradually in the transformation to B-mem^37^. rs2887399 is located upstream to *TCL1A*, the G allele is associated with both the higher risk of LOY^7^ and the upregulation of *TCL1A* in blood^38^. Thompson et al had spotted an overexpression of *TCL1A* in B-cells with LOY in comparison to non-LOY cells^9^. Here, our observation that LOY is overrepresented in B-in compared with B-mem provides a phenotypic explanation for the upregulation of *TCL1A* in B cells. On the other hand, *CD27* is a marker for B-mem cells. The G allele of rs10849448, upstream of *LTBR*, was significantly associated with higher risk of LOY ^9^. By exploring eqtlgene database of cis-eQTLs^38^, the G allele is associated with down regulation of *LTBR* (G allele, z= −29.8, *P*= 2.91×10^-195^), and interestingly the nearby *CD27* (G allele, z= −5.4, *P*= 7.1×10^-8^) ^38^.

A recent study has reported an exclusive expression of *TCL1A* in *TET2*- and *ASXL1*-mutant hematopoietic stem cells, and multipotent progenitors, normally these cells do not express *TCL1A*^39^. This discrepancy raises questions about relationship between *TCL1A* and LOY in B lymphocytes: does TCL1A drive LOY, or is LOY simply a marker of a phenotypic shift within the B-cell compartment? Our results collectively show that LOY is associated with bias toward B-in cells marked by the over expression of *TCL1A* opposed to CD27+ B-mem that lack *TCL1A*.

### Differential gene expression

To capture the expression changes in autosomal and X-chromosome genes associated with LOY, we assessed the differential gene expression between LOY cells, and non-LOY cells in each cell type (Supplementary Table 4).

*Genes encoding cell surface immunoproteins*: Previous studies have highlighted a downregulation of the surface protein *CD99* in LOY cells ^22^, we confirmed this observation in the majority of cell subtypes with the strongest effect in Treg (log_2_ FC= −0.67, *P_adj_* = 1.15×10^-31^), no effect in CD8nc, but with opposite direction in CTL (log_2_ FC= 0.17, *P_adj_* = 8.37×10^-26^). Although, *CD99* is a Pseudoautosomal region (PAR) gene, we observed the downregulation of other non-PAR cell surface markers; *CD48* located on chr 1 in all CD4+ T cells subtypes, and *CD40LG*, a gene located on the non-PAR of x chromsome, in Treg, and CD4et cells. Recently, reduced expression of *CD48* were detected in T cell lymphoma, and presented as causal for evading NK-cells dependent immunity^40^.

*Genes located on X-chromosome:* A recent study had detected an aberrant over expression of *XIST*, a long non-coding RNA located on X chromosome, in male cancers ^41^. *XIST* is essential for X chromosome inactivation (XCI) and normally expressed in female cells. We observed an over expression of *XIST* in LOY cells from all cell types. To assess the potential XCI features associated with LOY, we classified X-chromosome genes in our results analysis; following a previously curated list of the inactivation degree ^42^, into inactive, escape (genes escape XCI mechanism) and variable, and according to locus into PAR, and non-PAR. Within 99 genes classified as escape in non-PAR genes, we identified 26 genes (*XIST* + 25 genes) with DGE *p*_adj_< 0.05 in at least one cell type. The majority of these genes (23 out of 26) were significantly upregulated, with no cell type showing downregulation, whereas two genes showed both upregulation and down regulation, and one gene was only downregulated (Fig. 3a, Supplementary Table 4). Within each cell type, on average 37% of escape genes are upregulated in comparison to 4.8% downregulated (Wilcox rank test, *P*=0.04, results detailed in Supplementary Table 5).

**Fig. 3.**
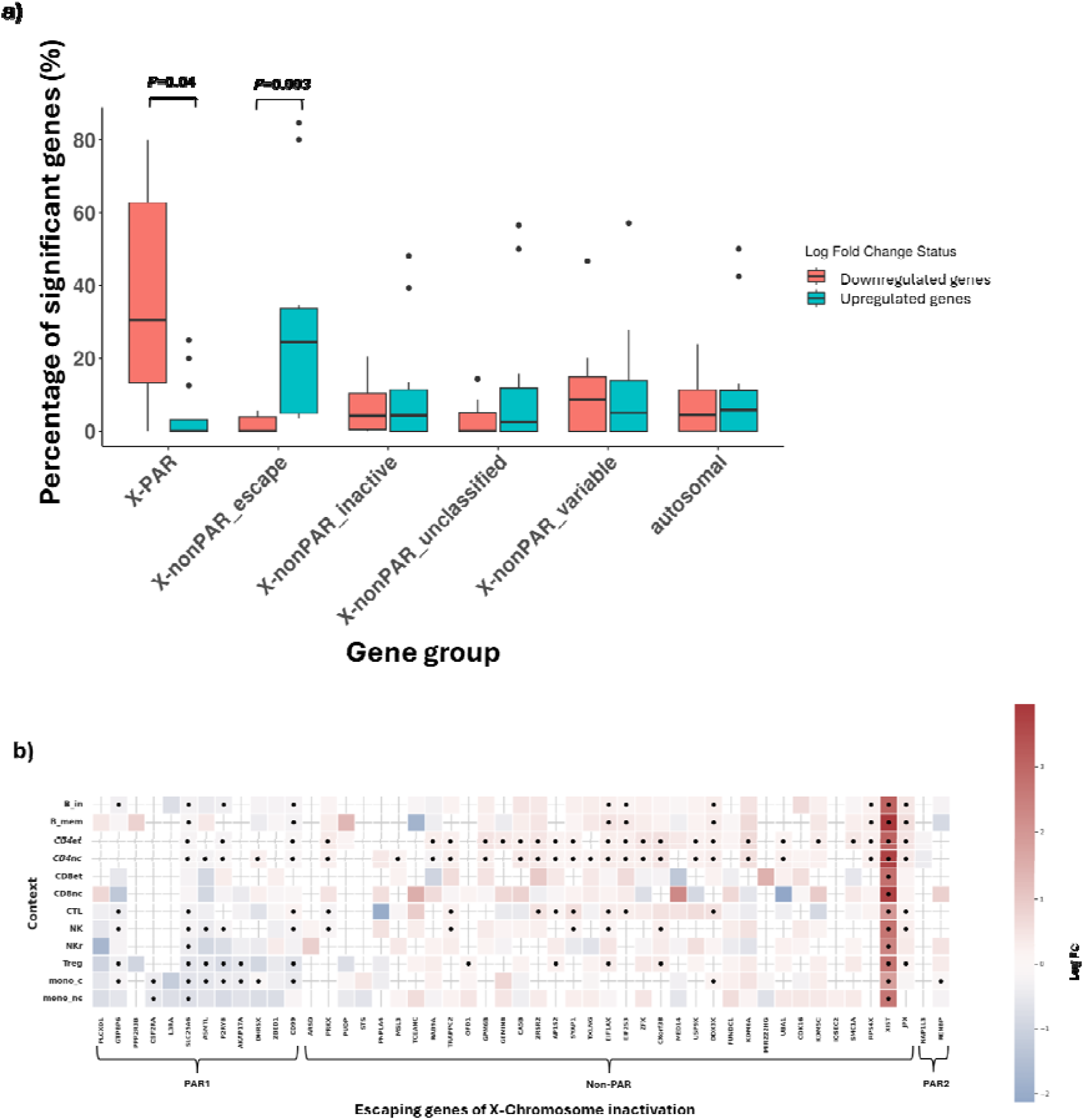
A combined representation of differentially expressed genes between LOY and non-LOY cells. (a) Box plot showing the percentage of significantly differentially expressed genes in each cell type (n=12). Genes are categorized based on their chromosomal origin: autosomal genes, X chromosome genes in the PAR region, and X chromosome genes in the non-PAR region. The non-PAR genes are further classified as escape, inactive, variable, or unclassified following Tukiainen et al., Nature (2017). Outliers are represented as black dots. (b) Heatmap displaying differentially expressed genes across 12 cell types, and focused on escape XCI genes. Colors represent log fold-change (logFC), and significant results (adjusted p-value < 0.05) are marked with dots.

To eliminate the possibility of contamination with female cells, we repeated the analysis after excluding cells that express *XIST* (Supplementary Table 6) and observed a similar trend that 19 out of 25 escape genes were significantly upregulated in all cell types. The 19 escape genes (Fig. 3B) include, *KDM6A*, *DDX3X*, and *KDM5C*. These three genes have been identified to harbour more loss of function mutations in male cancer samples in comparison to females in TCGA cohort ^43^. Similar male bias was identified in *ZRSR2*, a splicing gene associated with myeloid malignancies, and upregulated in T-cells with LOY ^44^. The 19 genes also include, *JPX*, a long non coding regulator for XCI, and *RPS4X*, a ribosomal gene that is over expressed in transformed myelodysplastic syndrome ^45^. *EIF1AX,* X-linked homolog of *EIF1AY*, that was identified, in addition to *DDX3X* , as the top gene expression dependencies for LOY cell line ^46^. On the other hand, only 23% of the inactive genes (n=143/431 genes) showed significant DGE in at least one cell type with a balanced distribution of association direction, between upregulated genes (n=74), and downregulated genes (n=43) in all cell types. Finally, nine X-PAR genes showed significant DGE between LOY and non-LOY (*P*=2.9×10^-3^), six genes were downregulated with non cell type upregulated (*P2RY8, SLC25A6, GTPBP6, ASMTL, CSF2RA,* and *AKAP17A*).

We explored the statistical results from a recent study that screened cell lines to detect gene dependencies of LOY cell lines in comparison to normal cell lines ^46^. 60 out of 17,385 genes showed significant differences (q value < 0.05) between LOY and wild type (WT) cell lines from cancer cell Line encyclopedia^47^ . Four escape XCI genes located on non-PAR (*RPS4X*, *DDX3X*, *ZFX*, and *EIF1AX* ) have the largest difference in average expression between LOY and WT cell lines, that refer to LOY cell lines dependency on these genes. These results collectively show XCI characteristics associated with LOY, in particular, the upregulation of genes escape XCI. These genes are known for sex-bias in healthy tissue ^42^, haematological and non-haematological cancers^41^.

### The dynamic effect of LOY on cells

To further understand the way that LOY affect immune cell fate, we carried two orthogonal analyses (i) We conducted gene enrichment analysis for LOY associated genes, and used hallmark gene sets from MSigDB ^48^. Genes were selected if they have logFC > 0.25 for upregulated genes, and < −0.25 for downregulated genes, and the analysis were restricted to cell types with the largest representation of LOY for each lineag (full enrichment results are presented in Supplementary Table 7) (ii) we assessed the effect of LOY on gene expression across five pseudotime trajectories of cell transition from B-in to B-mem, mono-c to mono-nc, NK to NKr, CD8nc to CD8et, and CD4nc to Treg. The analysis was restricted to genes that expressed in at least three of six quantiles using both a linear and quadratic model, and adjusting the analysis for age, and 10 PCs ^23^. these two approaches provide a functional view of the dynamics effect of LOY across different cell types.

Over the monocyte trajectory, LOY decreased the expression of non-classical monocytes markers *LYPD2*, and *C1QA*, and increase in classical monocytes markers *RNASE2*, and *VMO1*, confirming our previous lineage bias finding ^49^. Interestingly, the highest impact of the interaction between LOY and pseudo time was achieved on *IGFBP6* gene, an IGF binding-protein regulator, that increase monocytes migration from blood to tissue ^50^. Furthermore, the gene enrichment analysis of mono-c highlighted *MYC* transcriptional factor targets as the most significant pathway for the downregulated genes (Fig. 4a), however this did not include significant change in *MYC* gene expression, but we observed downregulation of *MYC* paralog: *MYCL*. Interestingly, *IL1B* was significantly downregulated in mon-c cells with LOY, and not in any other cell type. *IL1B* is a marker for inflammation, and its downregulation in macrophages suggests a bias toward fibrotic macrophages rather than inflammatory ^13^. Additionally, the downregulation of *LGALS2* in mono-c cells with LOY indicates the skewness of these monocytes to fibrotic phenotype. *LGALS2* encodes glactin-2, a CD14 ligand that induce the proinflammatory phenotype in monocytes and macrophages ^51^.

**Fig. 4:**
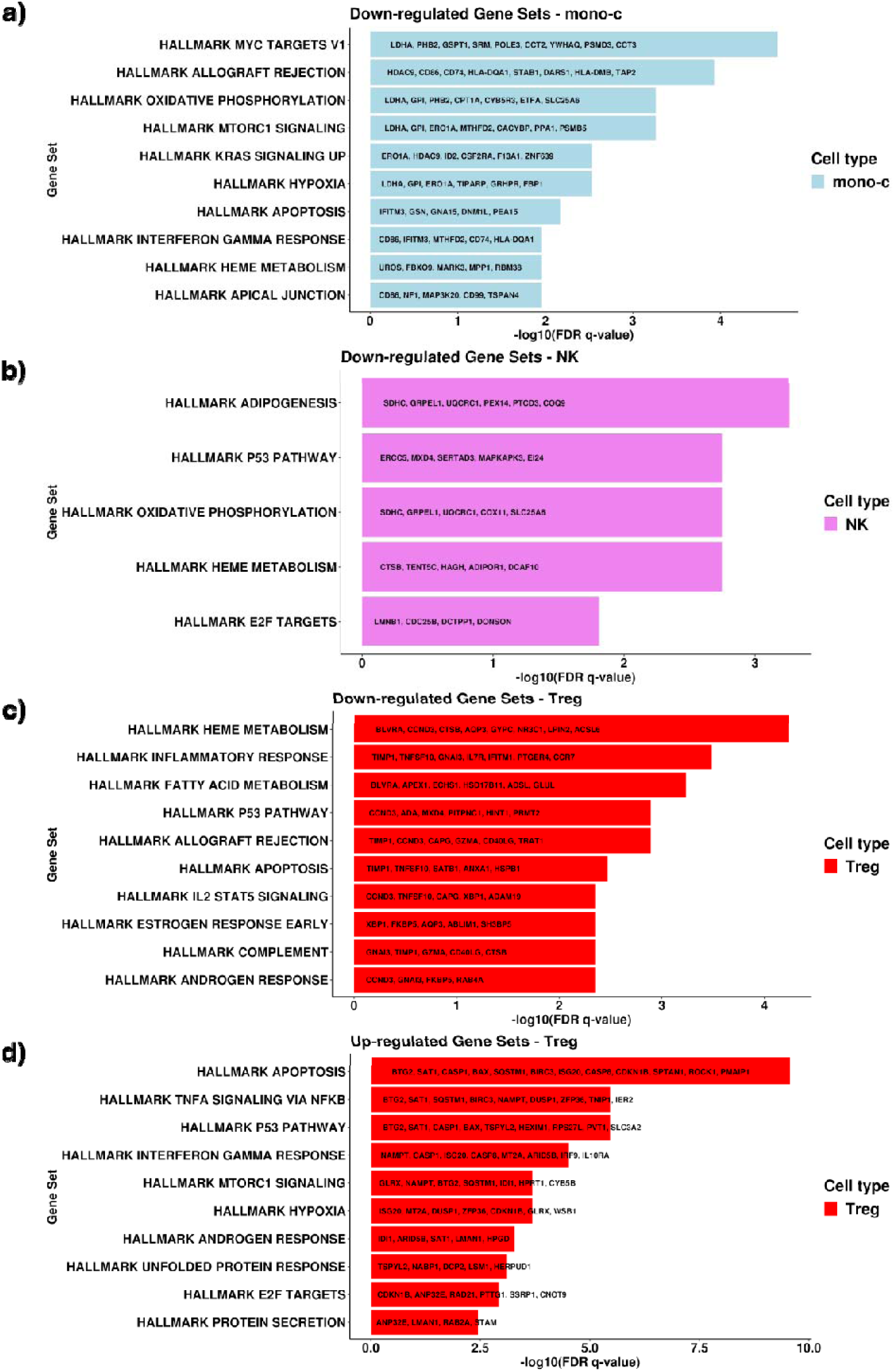
box plot of gene enrichment results using molecular signature data base( MSigDB), and focusing on significantly () differentially expressed genes between LOY cells and nonLOY cells in cell types with over representation in each pseudotime trajectory. This analysis were conducted on results excluding XIST expressing cells (a) down regulated genes with log FC < −0.25 in classic monocytes (b) down regulated genes with log FC < −0.25 in NK cells (c) down regulated genes with log FC < −0.25 in Treg (d) up regulated genes with log FC > 0.25 in Treg

NK cells show a significant decrease in *XCL1* associated with the LOY and the transition towards recruiting Natural killer cells (NKr). *XCL1* is a chemokine that is produced in activated NK cells, that supports our previous observation that LOY attenuates NK activation to NKr. The down regulated genes associated with LOY are notably enriched in heme metabolism and inflammatory markers (Fig. 4c). In contrast, upregulated genes are significantly overrepresented within the hallmarks of adiposis (Fig. 4e). These observations suggest that LOY may attenuate NK cell activation, potentially leading to alterations in NK function, particularly in the context of inflammation and metabolism.

In the trajectory of developing Treg cells, *FOXP3*, and *RTKN* have the most significant positive relationship with LOY, both are known markers for Treg cells (Supplementary Table 8). On the other hand, *ANXA1* is the most significant down regulated gene. These results confirm previous observations, that LOY polarizes CD4+ T cells toward a Treg state and induce an abnormal function ^19^. Furthermore, gene enrichment analysis of upregulated genes in Treg cells with LOY reveals a strong representation of apoptosis, p53 signalling, and the TNFα signalling via NF-κB pathway (Fig, 4e).

Finally, LOY exhibits its strongest effect in B cells by the downregulating of *HLA-A,* that reflects the bias toward B-in. HLA-A has previously connected to the risk of LOY in UK Biobank through GWAS ^9^. However, enrichment analysis did not identify any significantly associated hallmark gene sets in B cells or CD8+ T cells.

These results collectively, suggests that LOY alter the fate of immune cells, as exemplified by LOY monocytes being driven toward classic monocyte phenotypes with high affinity to migrate into tissue and transform into fibrotic macrophages.

## Discussion

Age-related somatic LOY in leukocytes is known to have a wide variation in the prevalence across cell types and is connected to different age-related diseases^17–19^. Here, we identified lineage specific biases across the developmental trajectories of the major cell types. Notably, we found a high prevalence of LOY in mono-c cells, resulting in expression profiles that suggest a transition towards fibrotic, rather than inflammatory, macrophages. Furthermore, we expanded the previous knowledge regarding the molecular effect of LOY^22^, on autosomal genes^17^ and methylation^21^, and included XCI characteristics, mainly upregulation of escape XCI genes, and aberrant expression of the long non coding *XIST*.

Our analysis captured a continuous drop in LOY cells across the trajectory of developing B cells, NK, and monocytes, and increase in LOY along the developmental trajectory of Treg, and CD8+ T cells. These trends expand on the previously observed over representation of LOY in monocytes, NK cells, and Treg. Variations across different cell types may be attributed, in part, to differences in cell lifespan. For instance, B cells have varying lifespans, ranging from weeks for naïve B cells to years for memory B cells, or longer for CD4+ T and CD8+ T cells. However, the shorter lifespans of monocytes (1-2 days), indicates a drastic impact of LOY on cell fate, that cell lifespan cannot explain.

Interestingly, the bias of LOY in B cells toward B-in marked with *TCL1A*, rather than B-mem marked with *CD27* suggests an explanation for the previous finding of overexpression of *TCL1A* in B-cells ^9^. Our finding shows that the overexpression of *TCL1A* in LOY cells is related to attenuating the development from B-in to B-mem. This supports the previous finding that *TCL1A* effect on clonal expansion is derived by its aberrant expression in haematopoietic stem cells^39^. rs2887399 near *TCL1A* was the first SNP to be connected to the risk of LOY ^7,9,15,21^. The bias of LOY toward CD27-B-in cells provides a potential fine-mapping for rs10849448, which has previously been associated with LOY^9,15^. This SNP is located in *LTBR* (12p13.31), and near *CD27.* While GWAS identified the association of G allele with higher risk of LOY, the study of eQTLs in blood connected it to the down regulation of *CD27* ^38^.

Remarkably, the bias of monocytes toward classical type with down regulation of pro-inflammatory cytokine, *IL1B*, has indicated possible pre-state for the migration of monocytes toward tissue and recruiting transformation to fibrotic macrophages, as classical monocytes, rather than nonclassical are known precursor for fibrotic macrophages ^52^. This observation expanded the previous finding that LOY induces the polarization of heart macrophages toward fibrotic macrophages (which lack *IL1B)* rather than inflammatory macrophages ^13^.

In addition, we showed for the first time a distinctive pattern of upregulation in escape XCI genes in comparison to other X-chromosome genes, and we reported an aberrant expression for *XIST* long noncoding RNA. These patterns had been identified in male cancer samples, both germ and non-germ lineages ^41^. Our findings support previous observation of the high dependency of LOY cell lines on a set of escape XCI genes with paralogues on Y chromosome (*RPS4X*, *DDX3X*, *ZFX*, and *EIF1AX*). This phenomenon could be explained by a compensation mechanism to protect the cell from LOY complications. While a small proportion of cells from female donors showed MSY mapped reads, that indicated the possibility of false negative in our LOY calls, but we prioritized specificity over sensitivity. Furthermore, the exclusion of cells with *XIST* expression did not affect the results.

These results demonstrate a wider effect of LOY on immune cells fate, triggering abnormal functions and mediating the effect on disease pathogenesis by; (i) In the myeloid compartment, LOY is associated with the transition of classical monocytes toward a profibrotic phenotype (ii) In the lymphoid compartment, LOY attenuates the activation of NK cells, and the transformation of B cells to B-mem, whereas it polarizes T cells toward Treg (iii) LOY is associated with aberrant expression of XCI-escape genes and XCI regulator *XIST.* Collectively, these insights establish LOY as a pivotal regulator of immune cell fate decisions, providing a compelling new framework to understand how age-related genomic instability shapes immune dysfunction and disease risk in clonal haematopoiesis, opening promising avenues for targeted diagnostics and therapeutic intervention.

## Methods

### Study cohort

OneK1K cohort was described in details elsewhere ^23^. In this study, we focused on males (n=416) for whom both scRNA-seq and genome-wide SNP arrays data are available. All donors in this subset are identified as healthy and age range between 19 and 93 years (median=68)

### Alignment, and counting of single-cell RNA-seq data

We obtained raw and multiplexed FASTQ files from Sequence Read Archive (SRA) for runs (SRR18028378 to SRR18029877) by using fastq-dump tool from NCBI SRA-tools. Next, we grouped FASTQ files according to analysis pools (n=75) and processed each pool independently using the default parameters of Cell Ranger (v8.0.1) ^26^, and GRCh38 reference release provided by Cell Ranger (refdata-gex-GRCh38-2024-A, March13, 2024). Cell Ranger generated a BAM file, and gene cell matrix for each pool. Next, the generated pool-level BAM files and gene cell matrices (counts) were demultiplexed using subset-bam (v1.1.0, https://github.com/10XGenomics/subset-bam ) and Seurat subset function ^53^, respectively, into donor-level files by following the published individual donor barcodes file obtained from Gene Expression Omnibus (GEO) under ID: GSE196830. Furthermore, we processed the donor-level BAM files using velocyto (v0.17) ^27^, that count and classify reads into spliced, unsliced, and ambiguous .

### Calling LOY in immune cells

We used all reads generated by both Cell Ranger and velocyto and mapped to Male Specific Region (MSR) of the Y chromosome (GRCh38, start: 2,781,480 , end: 56,887,902) to classify cells into LOY or non-LOY. LOY status is defined by the finding of no reads mapped to MSY region.

Calling both LOY and mCA using SNP array data:

We used MoChA, a BCFtools extension to call LOY and mCA using long-phased genotypes from genomic SNP array data ^28^. Initially, we obtained the genotyping IDAT pairs of files (green and red) from GEO accession: GSE196829. Next, we converted IDAT into GTC files, and to VCF files, sequentially using BCFtools plugin (https://github.com/freeseek/gtc2vcf ), Illumina manifest files (GSA-24v2-0_A1.bpm, and GSA-24v2-0_A1.csv), and the expected intensities file (GSA-24v2-0_A1_ClusterFile.egt). Next, the VCF file is phased independently using SHAPEIT5, and imputed using impute5. finally, MoChA calls chromosomal alterations, and the default filters were applied to classify events into LOY, LOX, mCA (CNG, CNL, and CNN) events.

### Identification of major cell populations

To get a benefit from the quality control and normalization steps applied by the author, we obtained feature cell matrix from CellxGene in “rds” format, compatible with Seurat (v4) R package and assigned our LOY calls into it. The original feature cell matrix contains 1,248,980 cells in 981 donors. We excluded a minimal number of cells (n=784 cells), that had not been called in our Cell Ranger assignment, and we cannot detect LOY state of them. The excluded cells affect 373 donors by excluding 1 to 4 cells in each (548 donor has no change, 226 with 1 cell excluded, 85 with 2 cells, 43 with 3 cells, 19 with 4 cells excluded). Initially, we used the predicted cell types by the query-reference cell map to group cells into five major groups, B-cells, monocytes, NK cells, CD4+ T-cells, and CD8+ T-cells. Other cell types, and outlier contaminated cells expressing CD34+ were excluded. Additionally, we used group specific contamination exclusion features; *CD4, CD3G/D/E*, and *CD8A/B* for B-cells, monocytes, and NKs; whereases *CD8A/B* were used for CD4 T-cells, and CD4, *CD3G/D/E* used for CD8 t-cells.

To achieve more purity, we analysed each cell type independently. Initially, we log-normalized counts of each subset with a scale factor of 10,000 and calculated the scaled expression of the most variable 500 features, and adjusted for percent of reads that map to mitochondria genome, and the sequencing pool. We clustered each subset using Louvain algorithm and the constructed KNN graph based on the Euclidean distance of 30 PCs. Next, we assessed the differential expression of the cell type markers by comparing each cluster to other clusters and using Wilcoxon Rank Sum test. Next, clusters that did not show a significant upregulation of the expected markers ^23^, were excluded.

### The relationship between LOY status and cell subtype

We tested the relationship between LOY and cell type using logistic regression model adjusted for age, sequencing pool, 10 PCs and multiple testing. We applied two different settings (i) cell types were binarized into 0/1 for B-in vs B-mem, NK vs NKr, Treg vs other CD4+ T cells, CD8nc vs CD8et, and mono-c vs mono-nc (ii) cells were classified according to their position on the pseudotime trajectory and classified into six equal quantiles. In details, we generated low dimensional embedding using potential of heat diffusion for affinity-based transition embedding (PHATE), the previously scaled expression data of 500 highly variable genes, and 10 PCs ^54^. Next, we calculated the pseudo time using slingshot ^55^, and PHATE embeddings. We specified the start and end cluster based on biological knowledge judgement. Next, we split the cells into six quantiles across the slingshot pseudotime curve. The frequency of LOY in each quantile was calculated and presented in percentages.

### Differential Gene expression

We tested the differential expression between LOY and non-LOY cells using MAST algorithm, a two-part hurdle model that was developed to deal with the highly frequent dropout, and the bimodal distribution of the single cell expression data ^56^. The algorithm was applied on the log normalized data and using the default parameters in Seurat (v5); restricting the analysis to genes expressed in at least 1% of the cells within each group and requiring a minimum log fold-change (LogFC) of 0.1 between clusters. The results were reported using average log2 fold change, percentage of cells where the gene is detected in, and Bonferroni adjusted p value. The differential gene expression analysis was replicated after excluding cells expressing XIST RNA.

### The dynamic effect of LOY on cell fate

To test the dynamic effect of LOY on gene expression. We adopted the method used in Yazar et al ^23^ to assess the linear and quadratic effect of the interaction between LOY and quantile, i.e the position of the cell on the trajectory. For each test, we generated two models: basic and augmented model. We compared the two models using anova test. The independent variables of the basic model are LOY status, quantiles of pseudotime, age, 10 PCs and under the random effect of donor. The augmented model includes the same variables in the basic model in addition to the interaction term between LOY status and quantiles of pseudotime. The independent variables of the quadratic model are LOY status, quantiles, quantiles square, age, 10 PCs and under the random effect of donor id. The augmented model includes the same variables in the basic model in addition to two interaction terms: LOY status and quantiles interaction, and LOY status and quantiles square interaction. For each gene, we reported the results of the linear and quadratic model as interaction estimate, p value, and Bonferroni adjusted p value.

### Gene enrichment analysis

We performed Gene Set Enrichment Analysis (GSEA) using hallmark gene sets from MSigDB to evaluate the enrichment of differentially expressed genes (DEGs). Genes were analysed separately for each cell type, with upregulated genes (logFC > 0.25, >0.5, and 0.75) and downregulated genes (logFC < −0.25, < −0.5, and < −0.75) assessed independently.

### Other statistical methods

The relationship between the percentage of LOY cells per donor and their age were tested using linear regression model in R. LOY percentages were logit-transformed to account for their bounded distribution between 0 and 100%. Age, 10 PCs and pool number were added as independent variables. The correlation between LOY clonal fraction estimated from SNP arrays, and that estimated from scRNA-seq, was tested using Pearsons’s correlation test. To compare the percentage of upregulated genes with the percentage of downregulated genes across different cell types, we categorized genes based on their chromosomal location into PAR, escape non-PAR, and inactive non-PAR groups. We then applied the Wilcoxon rank-sum test and reported the results as mean, median, p-value, and Wilcoxon statistic.

## Data Availability

All data produced in the present work are contained in the manuscript

https://github.com/ad2n15/LOY_OneK1K_Shiny

## Acknowledgement

A.D. and O.R acknowledge the support of this work from by Biotechnology and Biological Sciences Research Council [project reference: BB/Y513003/1]. L. G acknowledges Black Futures Scholarship at University of Southampton. Additionally, we acknowledge the use of the IRIDIS High Performance Computing Facility and associated support services at the University of Southampton.

## Data availability

The single cell raw sequencing DATA was obtained from SRA for runs (SRR18028378 to SRR18029877). The metadata and the genome variation profiling by SNP array were obtained from NCBI’s Gene Expression Omnibus (GEO) under ID: GSE196735, GSE196830, GSE196829. scRNA-seq counts were obtained from https://cellxgene.cziscience.com/datasets

LOY calls, cells clusters, and pseudotime inference can be visualized and downloaded from the shiny app. The app URL are available in https://github.com/ad2n15/LOY_OneK1K_Shiny

